# Dorsal root ganglia atrophy and serum biomarkers supporting the diagnosis of chronic postsurgical inguinal pain

**DOI:** 10.1101/2024.06.18.24309141

**Authors:** Eva Herrmann, Magnus Schindehütte, Gudrun Kindl, Ann-Kristin Reinhold, Felix Aulbach, Norman Rose, Johannes Dreiling, Daniel Schwarzkopf, Michael Meir, Yuying Jin, Karolin Teichmüller, Anna Widder, Robert Blum, Abdelrahman Sawalma, Nadine Cebulla, Michael Sendtner, Winfried Meissner, Alexander Brack, Mirko Pham, Claudia Sommer, Nicolas Schlegel, Heike L. Rittner

**Affiliations:** University Hospital Würzburg, Department of Anaesthesiology, Intensive Care, Emergency and Pain Medicine, Centre for Interdisciplinary Pain Medicine, 97080 Würzburg, Germany; University Hospital Würzburg, Department of Surgery I, 97080 Würzburg, Germany; University Hospital Würzburg, Department of Neurology, 97080 Würzburg, Germany; University Hospital Würzburg, Department of Neuroradiology, 97080 Würzburg, Germany; University Hospital Jena, Department of Anesthesiology and Intensive Care Medicine, Section Pain Therapy, 07747 Jena, Germany; University Hospital Würzburg, Institute for Neurobiology, 97080 Würzburg, Germany

**Author notes:** Corresponding author*: Heike Rittner, Centre for Interdisciplinary Pain Medicine, Department of Anaesthesiology, Intensive Care, Emergency and Pain Medicine, University Hospital Würzburg, Oberdürrbacher Strasse 6, D-97080 Würzburg, Germany. These authors contributed equally (Eva Herrmann, Magnus Schindehütte, Gudrun Kindl).

**Keywords:** Chronic postsurgical pain, hernia surgery, medication, imaging, cytokines, lipids

## Abstract

**Background:** Chronic postsurgical inguinal pain (CPIP) is the most common complication of groin hernia surgery. The characteristics of patients, their medical care, and choice of the best diagnostic tools remain to be defined to optimize preventive and therapeutic interventions.

**Methods:** Claims data from 2018 and a 1-year follow-up were analysed and deep phenotyping including sensory testing, blood and skin biopsies, MRI imaging of the dorsal root ganglion (DRG), and patient-reported outcomes were used to define normative values, as well as incidence, medical care, and pathophysiological factors.

**Results:** 11,221 patients with hernia surgery in 2018 were identified; 8.5% had pain before which was relieved by surgery, but a similar percentage had novel groin pain. Deep phenotyping of 141 healthy controls provided a map of the inguinal sensory system. CPIP patients suffered from moderate pain with neuropathic features, individual sensory abnormalities, and unilateral L1 DRG atrophy. In the blood, C-C-motif chemokine ligand (CCL2) and brain-derived neurotrophic factor (BDNF) were upregulated while apolipoprotein A1 (ApoA1) was reduced. A cluster of DRG atrophy, BDNF, ApoA1 and anxiety correlated best with the diagnosis. CPIP patients with novel pain had significantly more DRG atrophy (−22% ipsi vs. contra).

**Conclusion:** CPIP is relevant and often newly acquired after surgery. A combination of DRG imaging, serum markers, and anxiety screening can support the diagnosis. Using this core set of markers could guide surgeons towards more personalized therapies and possible preventive intraoperative techniques.

**Trial registration:** German Trial Registry DRKS00024588 and DRKS00016790

## Introduction

With more than 20 million operations annually, groin hernia repair is one of the most common surgical procedures worldwide.^1^ The development of chronic postsurgical inguinal pain (CPIP) is the most frequent problem in these patients with an incidence 10-54% depending on the criteria applied for the definition of CPIP.^2^ Despite technical advances in surgery, CPIP incidence has not declined in recent decades.^3, 4^

Chronic postsurgical pain newly develops or increases in intensity beyond the wound healing process ≥ 3 months after a surgical procedure.^3^ It is localized in the surgical area or its projection zones and often shows neuropathic pain characteristics. Known risk factors for chronic postsurgical pain include high levels of pre-surgical pain, severe postoperative pain, and psychological and other individual patient-related factors.^3, 5^ The exact pathophysiology of CPIP is not clear. 31-38% CPIP patients have a neuropathic component,^6, 7^ which is considered to be maintained by local and systemic inflammation.^8, 9^ A causative role of inflammation in the onset of CPIP has been assumed considering data from a small trial using anti-tumour necrosis factor-alpha (TNF-α) antibodies, which leads to reduced pain.^10^ Independent of the pain characteristics. All sensory stimuli of the groin are processed via the paired dorsal root ganglia (DRG) in the spinal cord segment L1. Whether local or systemic inflammatory changes, alterations in intraepidermal nerve fibre density (IENFD) at the site of surgery, or changes in the DRG itself occur in patients with CPIP remains unclear. The diagnosis of CPIP is dependent on clinical parameters that are subjective in nature. Accordingly, appropriate patient-tailored therapeutic approaches based on the pathophysiology or diagnostic tools of the CPIP phenotype are missing.

The present study was conducted to define the relevance of CPIP in real-world data, to use this knowledge to study the pathophysiology and possible neuropathy, and to identify additional diagnostic tools with deep phenotyping.

## Materials and Methods

A detailed description of the methods is found in the supplement.

### Analysis of the health care data (part of the LOPSTER project)

We used anonymized data from the scientific data warehouse of the German health insurance (BARMER GEK, DRKS00024588). Using the Operationen- und Prozedurenschlüssel (OPS) classification of procedures (OPS codes 5-530 or 5-531), we identified patients who underwent hernia surgery in 2018. Since CPIP is not coded in the ICD-10, surrogate diagnoses, specifically R10.2-4 (pelvic and perineal pain; pain with localization in other parts of the lower abdomen; other and unspecified abdominal pain) and M79.25 (neuralgia and neuritis, unspecified: pelvic region/thigh), were used before and after surgery. Sociodemographic and healthcare parameters were obtained one year before and after the index year 2018.

### Assessment of patient and control cohorts

The exploratory cross-sectional unicenter *ResolvePAIN* study protocol was approved (DRKS00016790). Seventeen patients with CPIP were recruited from the outpatient clinic of the Center for Interdisciplinary Pain Medicine or during a follow-up study at the Department of Surgery (**Supplementary Table 1**).^5^ Pain before surgery was assessed by telephone interview up to 1-4 years after the initial assessment. The control group comprised 141 cases subdivided into three age groups based on a sample size calculation using variability of pressure pain thresholds based on age and sex (**Supplementary Table 2**).^11^ All study participants underwent clinical assessment, patient-reported outcomes for pain and comorbidities, and quantitative sensory assessment (QST) of the groin and hand for comparison.

### Building a model to identify measures associate with CPIP as diagnostic tools

A model was created to identify relevant measures associated with CPIP using machine learning. With the chosen variables as predictors, a binary logistic regression model was built with diagnosis (CPIP vs. Control) as the outcome variable, with the Python package’ statsmodels’.^12^ Odds ratios and their confidence intervals were computed for each variable to determine the most important factors influencing diagnosis.

### Statistical analysis

Statistical analyses and data visualization were performed using GraphPad Prism (version 9.4.0, GraphPad Software, San Diego, California, USA), SPSS (SPSS statistics version 29.0.0, IBM SPSS Statistics for Windows, Armonk, NY: IBM Corp), and R for the claims data. The probability values of α < .05 were considered statistically significant.

## Results

### High incidence of CPIP and low treatment intensity based on health care data

Data from the BARMER health insurance database revealed that 11,221 patients had undergone unilateral inguinal hernia surgery in 2018. They were categorized into four groups: no pain before and after surgery (pain 0/0), pain before but not after surgery (pain 1/0; ***pain resolution***), no pain before but pain after surgery (pain 0/1; ***novel pain***), and pain before and after surgery (pain 1/1; ***pain persistence***) in the following 12 months. The latter two were labelled ***“probable CPIP”***. More than three-quarters of the patients did not complain of pain (pain 0/0) (**Figure 1A**). Apart from the anatomical repair of the hernia, 9.6% of patients specifically benefited because their presurgical groin pain resolved (pain 1/0). On the other hand, an equal number (8.5%) suffered from novel pain (pain 0/1). In total of “probable CPIP” was present in 13%: of these 30.9% had experienced no pain resolution despite the surgery. Men were affected more frequently than women were, but persistent pain (pain 1/1) was relatively more common in females.

**Figure 1.**
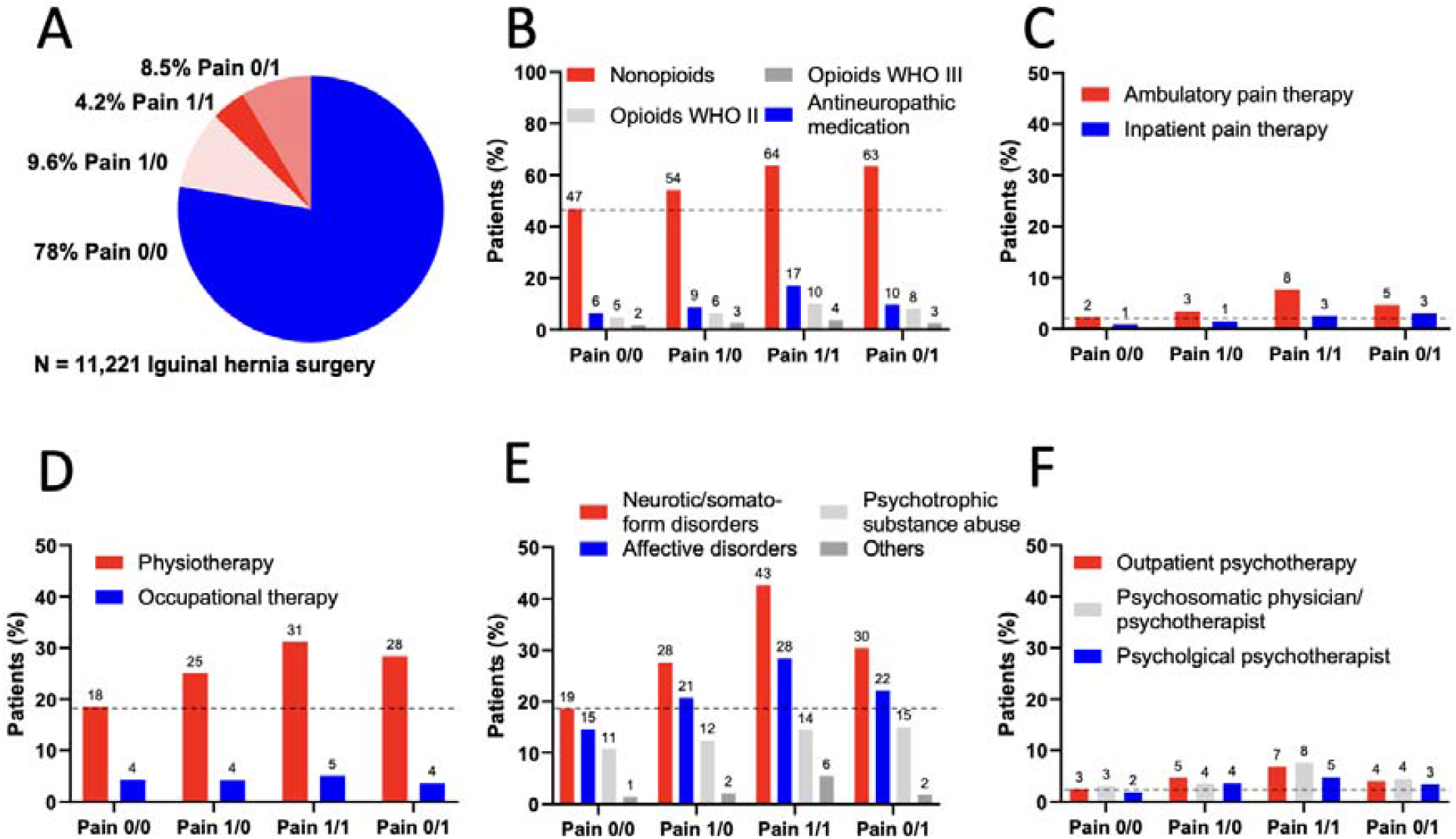
High incidence of “probable CPIP” after hernia surgery and treatment with nonopioids. Patients from a major German health insurance undergoing hernia repair in 2018 were classified by pain before and/or after hernia surgery using surrogate pain codes from the ICD-10. **A** Percentage of patients categorized in pain 0/0 (***no pain*** after surgery); Pain 1/0 (only pain before surgery = ***pain resolution***); Pain 1/1 (pain before and after surgery = ***pain persistence***) and Pain 0/1 (pain only after surgery = ***novel pain***). The latter two are labelled “probable CPIP”. Proportion of patients in “probable CPIP” subgroups receiving nonopioids, antineuropathic medication (anticonvulsants and antidepressants) or weak (WHO II) and strong (WHO III) opioids (**B**), specialized in- and outpatient pain therapy (**C**), physio- and occupational therapy (**D**), suffering from psychiatric comorbidities (**E**) and having psychotherapeutic treatment (**F**). n = 11,221.

Prescription of nonopioids of “probable CPIP” patients was not significantly different from patients without inguinal pain. Treatment with antineuropathic drugs, such as antidepressants and anticonvulsants, was highest in the pain persistence group (2.8 ×), followed by the novel pain group (1.6 ×) (**Figure 1B**). Patients with “probable CPIP” were rarely cared for by a pain specialist, either ambulatory (5-8%) or inpatient (3%) (**Figure 1C**). Physical and occupational therapies were also prescribed most frequently in the persistent pain group (**Figure 1D**). Psychiatric comorbidities, especially somatoform and affective disorders, were more prevalent but not necessarily more common in “probable CPIP” (**Figure 1E, F**). In summary, persistent pain and novel pain after hernia surgery are frequent but of these “probable CPIP” patients a maximum of one fifth receives treatment for chronic pain.

### Sensory phenotyping of the groin in healthy individuals

To understand the pathophysiology and clarify the neuropathic components of CPIP, we deeply phenotyped a selected group of patients. Because of the lack of normative data, we first established age- and sex-dependent reference data for sensory testing^13^ and skin innervation (**Supplementary Table 1**).^14^ Healthy participants (n = 141) had no general defect in sensation as thresholds of the hand were normal when compared with reference data from the German Research Network of Neuropathic Pain (**Supplementary Figure 1**).^11^ In the groin, only the deep pressure pain threshold but none of the other ten temperature and mechanical parameters showed a clear dependence on age and sex: male controls were generally less sensitive and thresholds further increased with age (**Figure 2A-K**). Deep pain thresholds were independent of BMI, excluding variances in subcutaneous fat (**Supplementary Table 2**).

**Figure 2.**
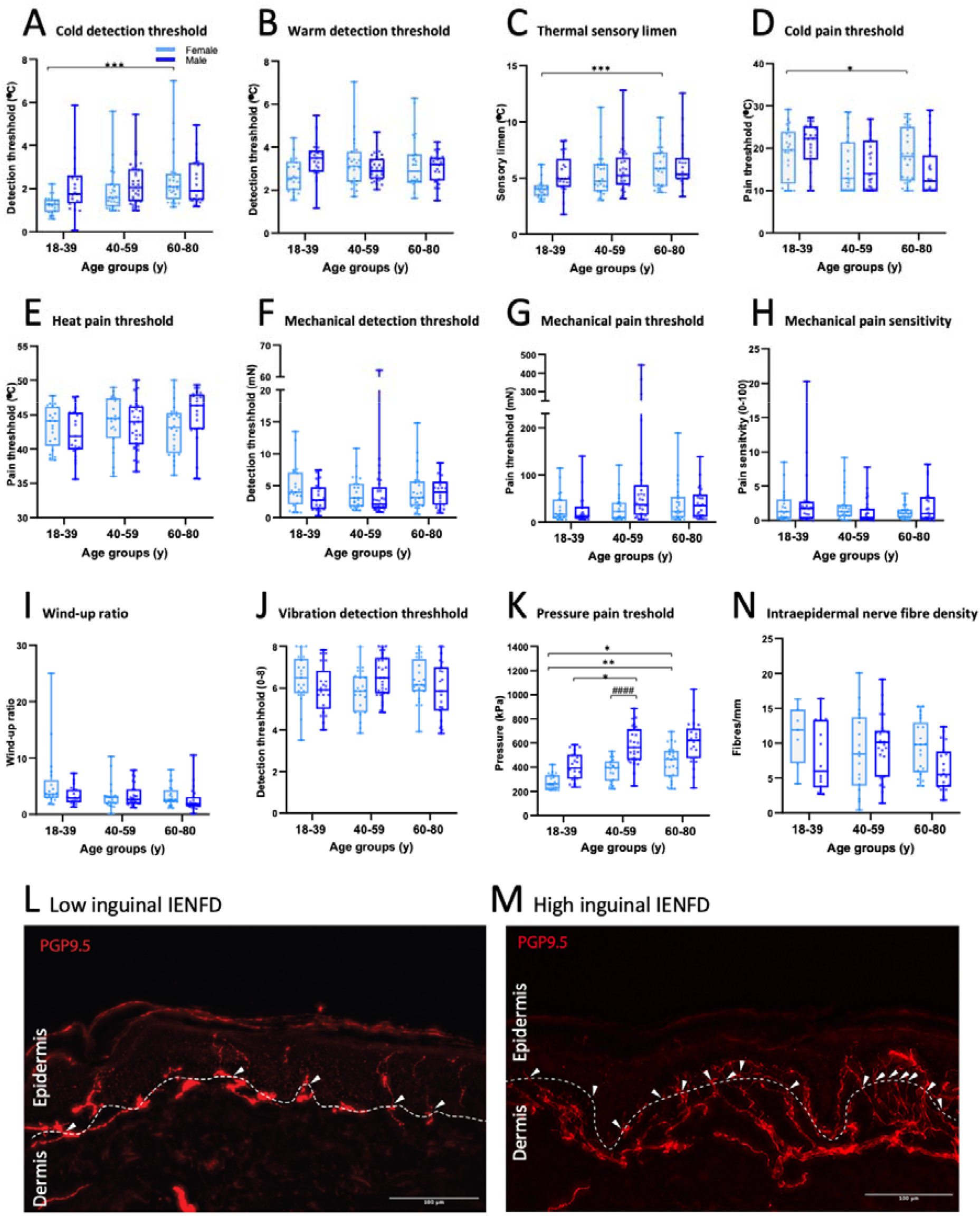
In quantitative sensory testing (QST), pressure pain in the groin is age- and sex-dependent. Healthy controls underwent QST of the groin as well as skin biopsy. **A** Cold detection threshold; **B** warm detection threshold; **C** thermal sensory limen; **D** cold pain threshold; **E** heat pain threshold; **F** mechanical detection threshold; **G** mechanical pain threshold; **H** mechanical pain sensitivity; **I** wind-up ratio; **J** vibration detection threshold; and **K** pressure pain threshold are displayed. **L** Skin punch biopsies were labelled with anti-PGP9.5. Representative examples with many (upper panel) and few fibers (lower panel) are depicted. **M** Normal values from the intraepidermal nerve fiber density (IENFD). Data are shown by age groups in males and females and presented as median, 25^th^ and 75^th^ percentiles on linear Y-axes. A-K: n = 141; M, L: n = 104. One way ANONA on ranks (Kruskal-Wallis test). Star symbols depict significant effects of age within one sex, hashtags show significant gender effects within one age group as follows: */#, p < 0.05; **/##, p < 0.01; ***/###, p < 0.001; ****/####, p < 0.0001.

Free nerve endings in the epidermis are responsible for pain transmission. This innervation is age- and sex-dependent in the distal leg.^14^ In general, groin skin biopsies from healthy controls had a high variance in IENFD with medians between 6-12.5/mm^2^, comparable to those previously described for the upper thigh (**Figure 2L, M**).^15^ Sex and age did not affect IENFD in this region.

### Selected nerve injury and overall DRG atrophy combined with elevated monocytic chemokine, and neurotrophic factors and lower anti-inflammatory lipids in CPIP

Equipped with normative data, 17 patients with CPIP were phenotyped to identify biological correlates. All available clinical tests, including patient-reported outcomes, sensory phenotyping, MRI-based imaging, and quantification of serum biomarkers were performed (**Figure 3A**). The basic characteristics of male and female patients with CPIP did not differ, but more female patients underwent open surgery between 4 months and 15.3 years after hernia repair (**Supplementary Table 3**). All CPIP patients experienced moderate mean pain, severe maximum pain, and low disability. Pain had some symptoms of neuropathic origin based on the Neuropathic Pain Symptom Inventory (NPSI). One-third of patients experienced mild symptoms of depression or anxiety. Most patients were taking nonopioids, and only a few were taking antineuropathic medication. In the analyses of the somatosensory system, thresholds for noxious and non-noxious stimuli in patients with CPIP were mainly within the normal range, based on Z-values calculated based on healthy controls (**Figure 3A, B**). Ten patients reported allodynia (**Figure 3C**) and/or thermal loss. In 11 patients, bilateral skin biopsies were available, 63% had a lower ipsi-to contralateral IENFD ratio (**Figure 3D**). Therefore, according to sensory testing, a subgroup of patients had signs of nerve injury; however, both tests did not necessarily overlap.

**Figure 3.**
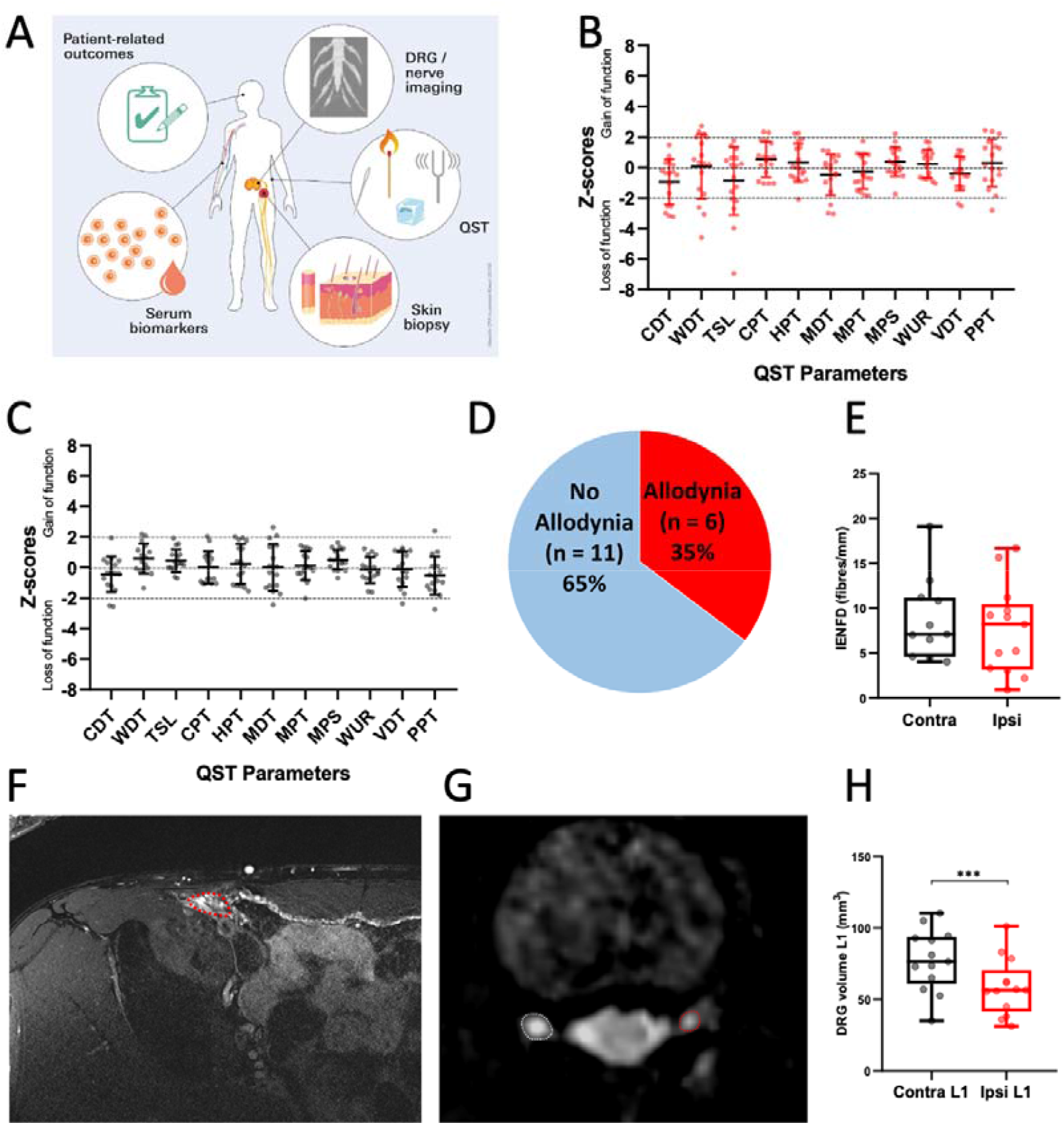
Unaltered sensory profiles and skin innervation in CPIP patients but ipsilateral L1 DRG atrophy. **A** CPIP patients from the cross-sectional study underwent deep phenotyping including QST, skin punch biopsies in the groin area, halfway between the iliac spine and the superior ramus of the public bone, as well as MRI neurography. **B, C** Ipsi- and contralateral sensory thresholds. Dotted lines depict two standard deviations. **D** Fraction of CPIP patients with mechanical allodynia. **E** Sensory fibers were labelled with PGP9.5 to calculate the intraepidermal nerve fiber density (IENFD) on the ipsilateral and contralateral side. **F-H** Axial T2-weighted MRI of the groin and bilateral DRG L1 after right-sided hernia repair. **F** Postoperative changes at the level of the inguinal canal (dotted red line) as well as the mesh (arrowheads). **G** Comparison of DRG level L1 of a patient with CPIP on the left side (ipsilateral DRG; red) compared to the contralateral unaffected DRG (white). **H** Evaluation of the DRG volumes of the inguinal dermatome (L1). The boxplot depicts median, 25^th^ and 75^th^ percentile. Wilcoxon test; ***p = 0.0002 (n = 17 of these 13 MRI and 11 skin biopsy). CDT, cold detection threshold; WDT, warm detection threshold; TSL, thermal sensory limen; HPT, heat pain threshold; CPT, cold pain threshold; MDT, mechanical detection threshold; MPS, mechanical pain sensitivity; MPT, mechanical pain threshold; WUR, wind-up ratio; VDT, vibration detection threshold.

On MRIs of the groin and its innervating structures, inguinal nerves were identified in some, but not all, patients (**Figure 3E**), probably due to limited spatial resolution and movement artifacts. Potential injuries to the ilioinguinal nerve or ramus genitalis of the genitofemoral nerve cannot be excluded. The surgically inserted mesh could be identified medially adjacent to the nerves in only some cases because of movement artifacts. No local inflammatory changes, usually revealed by tissue edema or contrast agent uptake during intravenous injection as the cause of groin pain, were observed in any of the images. The somas of the ilioinguinal and genitofemoral nerves were located in the bilateral L1 dorsal root ganglion (DRG), representing the inguinal dermatome (**Figure 3F**). Surprisingly, in the voxel-wise MRI DRG morphometry, DRG volumes of the affected side were reduced by 24.4% compared to the healthy contralateral side (n = 13, **Figure 3G**). In fact, a reduction in DRG volume was observed in all patients.

Of the nine cytokines linked to inflammation-induced neuropathic pain syndromes^9^, only CCL2 was significantly increased in CPIP patients compared with sex- and age-matched controls (**Figure 4A**). All other cytokines (IL-4, -6, -8, -10, -18, -27, TNF-α, and VEGF) were heterogeneous in patients compared with a more uniform control group (**Supplementary Figure 2**). Among the lipids, total HDL and ApoA1 levels were significantly reduced (**Figure 4B**), while cholesterol, triglycerides, LDL, and ApoA2 were unchanged. Brain-derived neurotrophic factor (BDNF) levels were higher in patients with CPIP (**Figure 4C**). The axonal damage marker NfL did not indicate overall group differences (**Figure 4D**).

**Figure 4.**
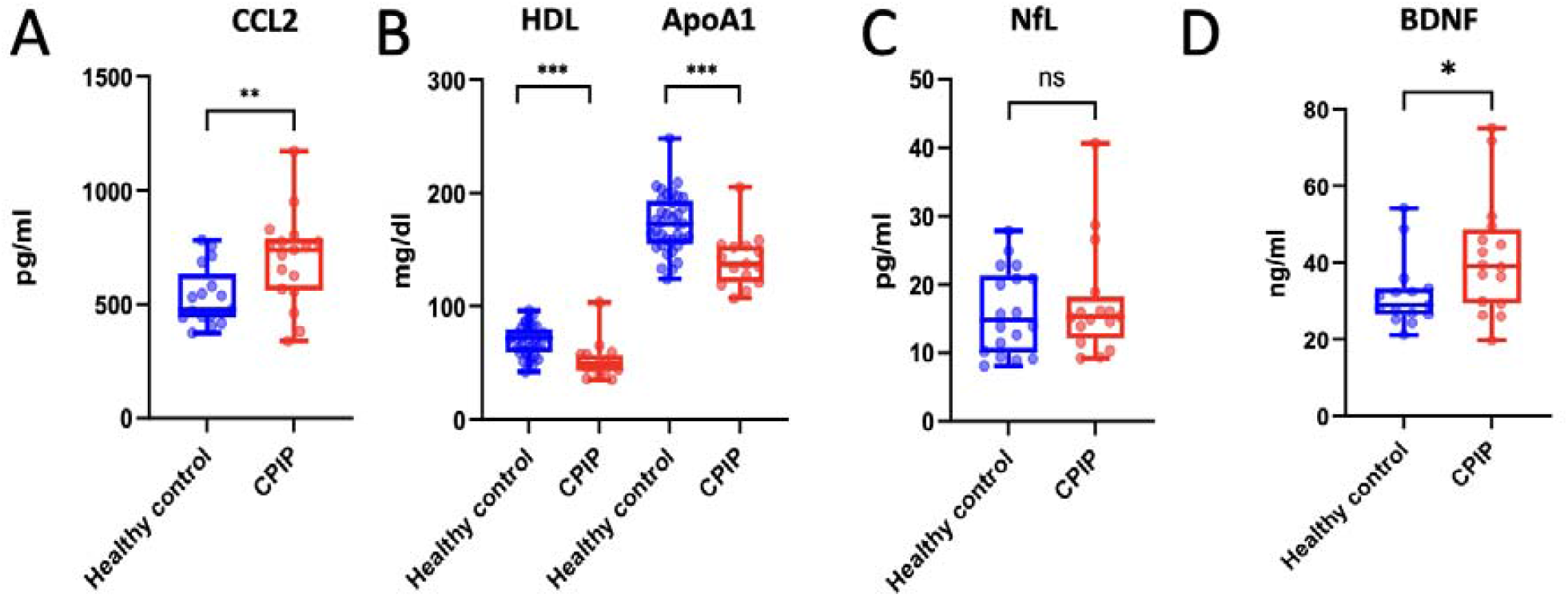
Increase in CCL2 and brain-derived neurotrophic factor (BDNF) but decrease in HDL and apolipoprotein A1 in CPIP. Nine blood cytokines, chemokines, lipid profiles, and two neuronal markers were measured in CPIP patients and compared to age- and sex-matched controls. **A** CCL2 **B** Lipid markers HDL and ApoA1 **C** Neurofilament light chain (NfL). **D** BDNF levels in the serum. n = 15-17, t-test or Whitney-Mann test, with Bonferroni correction for multiple testing. ** p < 0.05, *** p < 0.001, ns = not significant.

### More DRG atrophy in patients novel groin pain after surgery

Since health insurance data demonstrated that 69.7% of patients with “probable CPIP” had novel postsurgical pain, we subgrouped the data from the cross-sectional study. The patients’ baseline characteristics did not differ between those with or without presurgical pain (**Supplementary Table 4**). Interestingly, the ipsilateral L1 DRG volume of patients without presurgical pain was even smaller, pointing towards DRG atrophy, possibly due to nerve injury (**Figure 5A**). We further compared the two studies regarding treatment: drug treatment in CPIP patients from the cross-sectional study was similar to “probable CPIP” patients the health insurance cohort (nonopioids: 58.8% CPIP vs. 63-64% “probable CPIP”; opioids: 18% CPIP vs. 11-14% “probable CPIP”; antineuropathic drugs: 18% CPIP vs. 10-17% “probable CPIP”). Finally, we evaluated whether routine treatment matched already with the type of pain (inflammatory versus neuropathic); anti-inflammatory drugs were slightly more frequently used by patients with elevated CCL2 levels (60% in patients with high vs. 42% with low CCL2). Antineuropathic drugs were rarely prescribed (18% of patients) – and not more frequently prescribed in patients with allodynia.

**Figure 5.**
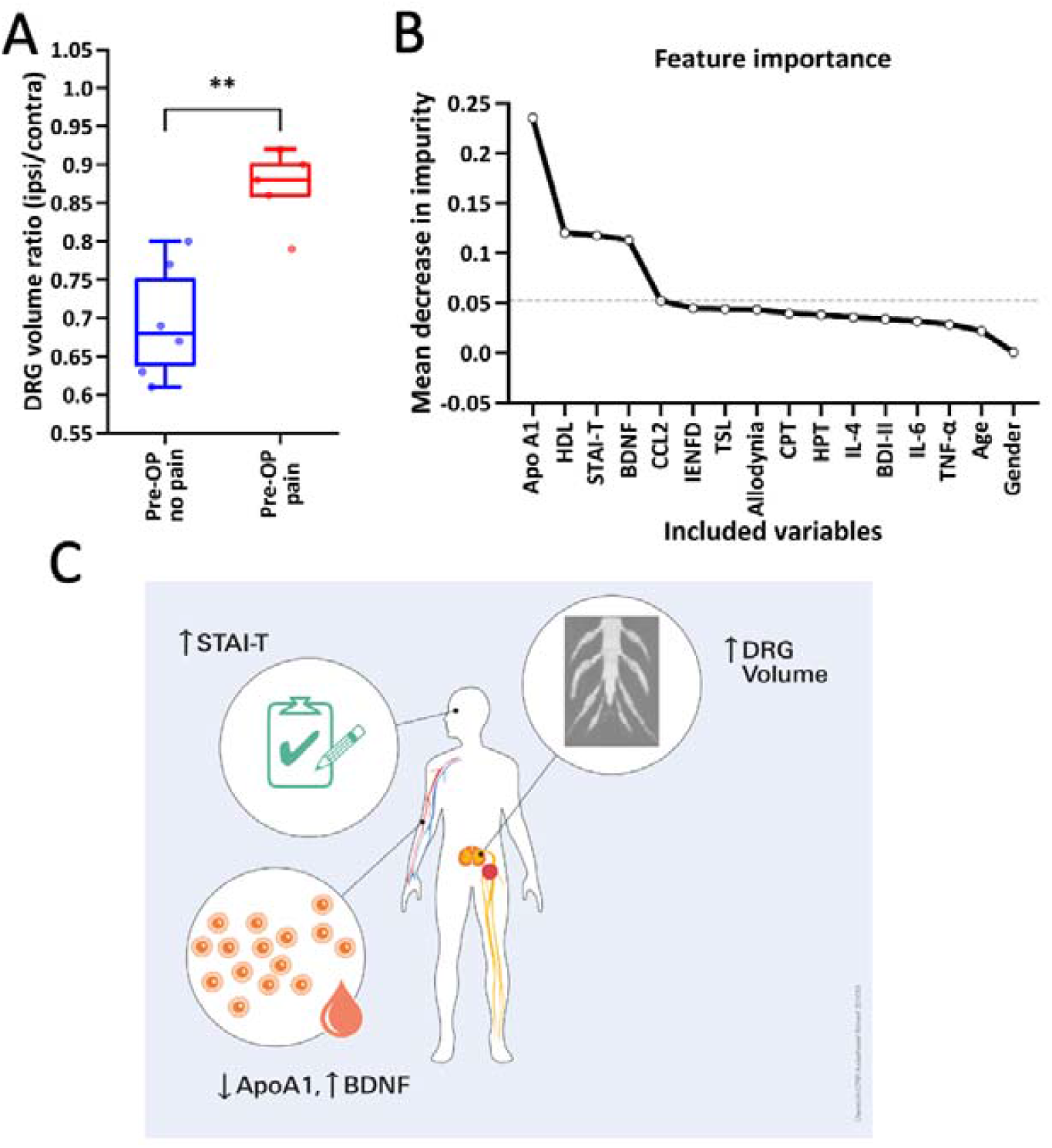
Diagnostic tools and different pathophysiology depending on the presurgical pain. **A** Comparison of the DRG volume level L1 between patients with and without presurgical pain. **B** Mean decrease in impurity was used to compute the most relevant factors. **C** Summary of suggested diagnostic tools. n = 11, t-test. ** p < 0.05.

### Low anti-inflammatory lipid proteins and high levels neuronal growth factors and anxiety as diagnostic tools

To identify the most important diagnostic parameters and to better understand the pathophysiology of CPIP apart from DRG atrophy, calculation of mean decrease impurity (MDI) was performed in 17 patients with CPIP and 15 healthy controls (**Figure 5B, C**). MDI depicted that BDNF, HDL, ApoA1, CCL2 and STAI-T were of the highest importance for the model. To test this hypothesis, we included four variables in the logistic regression model. The inclusion of HDL resulted in a VIF scores > 4.8 because to its high correlation with ApoA1. Therefore, they were removed from the analysis. The logistic regression model with ApoA1, BDNF, and STAI-T was significant, with a log-likelihood ratio p-value < 0.001 (**Supplementary Table 5**). The model showed that low ApoA1, high BDNF, and high STAI-T scores were associated with a higher probability of being in the CPIP group.

## Discussion

Many patients worldwide undergo elective hernia repairs. Our study confirms that every 10^th^ patient suffers from persistent inguinal pain or develops novel pain that is not present before surgery. There was a gap between diagnosed chronic inguinal pain (“probable CPIP”) and its proper treatment, for example, interdisciplinary multimodal therapy or antineuropathic medication, according to health insurance data. Analysis of a cohort of patients with CPIP revealed moderate to high pain intensity with low to moderate disability. Deep phenotyping based on the pathophysiology of nerve damage and persistent low-grade inflammation identified ipsilateral DRG atrophy, in combination with low ApoA1, elevated BDNF, and high anxiety levels, as the best additional diagnostic tools. Ipsilateral DRG atrophy is specifically pronounced in patients with novel pain.

Using healthcare claims data, we confirmed that approximately 12% of patients suffer from “probable CPIP” (groin pain) after hernia repair, in line with previous prospective^16^ and retrospective data,^5^ but in sharp contrast to other data (54%).^2^ While pain in some patients resolves with hernia repair, 30% (= 4% or 800,00 each year worldwide) permanently suffer from it.^16^ Pain impact on daily life is variable. The treatment of “probable CPIP” in our cohort consisted of standard non opioids after surgery but often lacked antineuropathic medication. In another study, only 2.87% of patients received a new prescription for analgesics. However, this study underestimated the incidence because it excluded preoperative analgesic prescription, which is a known risk factor.^17^ Our patients with “probable CPIP” had more psychosocial stress factors, including psychological distress, anxiety, catastrophizing, reduced pain coping, depression, and hypervigilance are known patient factors.^3, 18^ Whether resurgery is helpful or even harmful is not completely clear. Stratification based on pain sensitivity to pressure algometry for re-surgery and neuropathic pain for pharmacotherapy led to a better improvement in the re-surgery group.^19^ So there is room for improvement using biomarker-targeted treatments.

One option for markers is sensory testing, which can identify different pathological subtypes in patients with neuropathies of different origins.^13, 20-22^ Using healthy controls to generate normative data, we were able to thoroughly map the patients with CPIP. None of the CPIP patients had abnormal values in all QST measurements; changes were more subtle than those in other diseases.^20^ QST as a biomarker has limitations, not only in CPIP; as a psychophysiological method measuring thresholds, it does not map clinically relevant spontaneous and deep pain in CPIP.^23-25^ Since we identified allodynia in the clinical examination, which can be easily studied using a brush, QST seems dispensable. Suprathreshold stimulation or new assays for deep fibres will be useful in the future.^23^ Secondly, IENFD was not altered in CPIP patients.^22^ Possibly studying nociceptors themselves by labelling transient receptor potential vanilloid 1 (TRPV1) or calcitonin gene-related peptide (CGRP) will be better. In summary, we cannot recommend QST or skin biopsy at this stage.

In our panel, CCL2, a proinflammatory chemoattractant for monocytes, was increased, as previous seen in preclinical neuropathic pain models^26, 27^ and patients with polyneuropathy.^9^ In individual patients, TNF-α, IL-4, and IL-6 levels were elevated. Larger groups of patients are needed to better understand which subgroups of patients have these elevated cytokines. Among lipids, HDL, with its backbone ApoA1, which is mainly known to protect against cardiovascular diseases^28^ –, was reduced. Rare mutations of ApoA1 are associated with polyneuropathy (Tangier disease).^29^ Lipids are crucial players in inflammation. For instance, oxidized phospholipids are proalgesic and can be captured by the ApoA1 mimetic D-4F, thus preventing nociceptor activation in acute and chronic inflammatory pain.^30^ The second useful blood biomarker is BDNF, a neurotrophin involved in central sensitization.^31, 32^ BDNF increases nociceptor excitability in the DRG and the dorsal horn of the spinal cord.^33^ BDNF levels are also increased in fibromyalgia and other primary chronic pain disorders. ^34^ In contrast, NfL, a marker of axonal damage, was unchanged in CPIP.^35, 36^ In summary, our data indicate that CPIP has inflammatory and central nervous system remodelling features.

In high-resolution 3D DRG imaging, CPIP was associated with atrophy of the affected L1 DRG in all patients and was even more pronounced in patients with novel inguinal pain after surgery. On possible explanation is the neuronal loss in the outer layer of the DRG, as observed in neuropathy in diabetes^37^ or plexus injury.^38^ However data from preclinical models are controversial whether nerve injury leads to neuronal loss.^39^ Primary injury to the ilioinguinal or genitofemoral nerve or postsurgical compression or inflammation could be causative.^2, 40^ Direct detection of peripheral nerve injury of the genitofemoral or ilioinguinal nerve on MRI was not possible due to motion artifacts and limited spatial resolution. Further DRG imaging techniques such as microstructural or quantitative, functional MRI will be valuable in the future.

To test the predictive power of deep phenotyping, in addition to DRG atrophy, an AI-based model was developed that included all measures. After excluding irrelevant variables, ApoA1, BDNF, and the STAI-T were identified as the best predictors and biomarkers for CPIP. Lower levels of ApoA1 and higher levels of BDNF and STAI-T were associated with a higher probability of CPIP in our sample. However, our model is not intended as a substitute for clinical diagnosis. We recommend for futher validation in future patient cohorts.

A major limitation of healthcare data is the lack of an ICD-10 code for CPIP and no repeated pain measurements. Claims data are designed for reimbursement and might therefore be biased. Second, we did not know the cause of analgesic prescriptions, such as inguinal pain or other diseases. Our cross-sectional clinical study was limited by the small number of participants.

In summary, we provide data indicating that many patients who undergo elective hernia surgery suffer from novel groin pain. Therefore, patients scheduled for hernia surgery should be informed of this potential complication. Second, based on our exploratory study, we provide clinicians with additional diagnostic tools for CPIP, including DRG MRI, ApoA1 and BDNF serum levels, and anxiety screening with the STAI test. Together, they suggested more nerve-sparing techniques and individual therapies based on a core set of markers.

## Data Availability

All data produced in the present study are available upon reasonable request to the authors.

## Acknowledgement

We are grateful to all patients and healthy volunteers, as well as staff, especially Sandra Dinies (Centre for clinical surgical research), who contributed to this study.

## Authors’ contributions

Eva Herrmann: acquisition, analysis, and interpretation of clinical, QST and skin data, drafting the article, final approval. Magnus Schindehütte: acquisition, analysis and interpretation of MRI data, drafting the article, final approval. Gudrun Kindl: conception and design, acquisition, analysis and interpretation of claims data, drafting the article, final approval. Ann-Kristin Reinhold: acquisition, analysis and interpretation of blood biomarker data, drafting the article, final approval. Felix Aulbach: acquisition, analysis and interpretation of claims data, drafting the article, final approval. Norman Rose: acquisition, analysis and interpretation of claims data, drafting the article, final approval. Johannes Dreiling: conception and design of claims data, drafting the article, final approval. Daniel Schwarzkopf: conception and design of claims data, drafting the article, final approval. Michael Meir: analysis and interpretation of patient data, drafting the article, final approval. Yuying Jin: acquisition, analysis, and interpretation of skin data, drafting the article, final approval. Karolin Teichmüller: acquisition, analysis, and interpretation of claims data, drafting the article, final approval. Anna Widder: analysis and interpretation of patient data, drafting the article, final approval. Robert Blum: conception and design, critically revising the article, final approval. Nadine Cebulla: acquisition, analysis, and interpretation of blood biomarker data, drafting the article, final approval. Abdulrahman Sawalma: statistical analysis and interpretation of data, drafting the article, final approval. Michael Sendtner: analysis and interpretation of biomarker data, drafting the article, final approval. Winfried Meissner: conception and design, critically revising the article, final approval. Alexander Brack: conception and design, critically revising the article, final approval. Mirko Pham: conception and design, critically revising the article, final approval. Claudia Sommer: conception and design, critically revising the article, final approval. Nicolas Schlegel: conception and design, critically revising the article, final approval. Heike L. Rittner: conception and design, writing and critically revising the article, final approval.

