## Supplemental Figures for "Dorsal root ganglia atrophy and serum biomarkers supporting the diagnosis of chronic postsurgical inguinal pain"

### Supplementary Figure 1

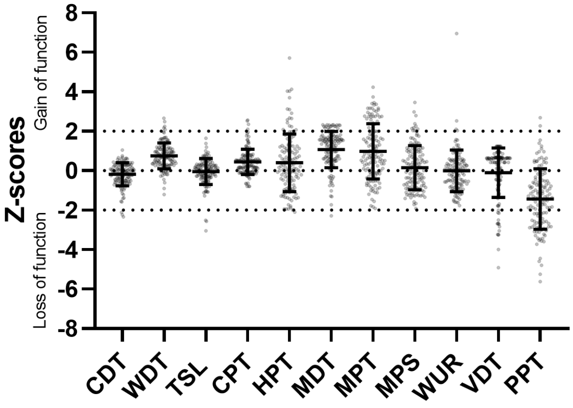


**Supplementary Figure 1: Control quantitative sensory testing values of the right hand in healthy controls.** Cold detection threshold (CDT), warm detection threshold (WDT), thermal sensory limen TSL), cold pain threshold (CPT), heat pain threshold (HPT), mechanical detection threshold (MDT), mechanical pain threshold (MPT), mechanical pain sensitivity (MPT), wind-up ratio (WUR), vibration detection threshold (VDT), and pressure pain threshold (PPT) are displayed.  N = 141 healthy controls. Dotted lines delineate the range of two standard deviations.

### Supplemental Figure 2

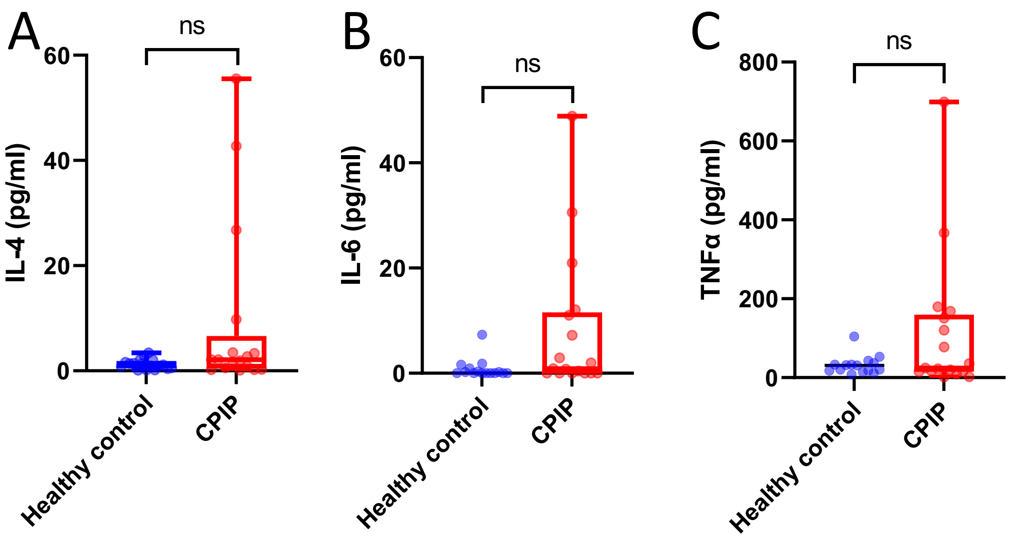


**Supplementary Figure 3: No difference in other pro- and anti-inflammatory cytokines.** Blood cytokines were measured in CPIP patients compared to age- and sex-matched controls. **A** Interleukin (IL)-4 **B** IL-6 **C** Tumor necrosis factor (TNF)$\alpha$. n = 15-17, t-test or Whitney-Mann test, with Bonferroni correction for multiple testing. ns = not significant.
