## Supplemental Information for "Dorsal root ganglia atrophy and serum biomarkers supporting the diagnosis of chronic postsurgical inguinal pain"

### Supplementary information

### Materials and Methods

#### *Analysis of the health care data*

We used anonymized nationwide administrative claims data warehouse for inpatient and outpatient treatment as well as core data of a German administrative health care claims data (BARMER GEK). In Germany, the vast majority of workers, along with their families, are covered by public health insurances. Public insurances provides comprehensive coverage, including hospital and outpatient care, medications, and other therapeutic aids. 90% of the population in Germany are covered by public insurances. So, these data are quite reflective of the broader German demographic. Various public health insurance schemes exist, such as BARMER, offering similar benefits and collecting comparable data. Individuals are free to choose among different public health care plans. Minor variations exist among different insurance plan members due to historical factors; for example, BARMER's enrollees include a higher proportion of women, yet it still offers a good representation of Germany's population demographics (approximately 9 Mio insured persons, 10% of total German population).[1] In terms of medications, the German claims database records every prescription issued by doctors for outpatient care under each health insurance scheme, but lacks data on in-hospital prescriptions. In summary, we accessed outpatient opioid prescriptions from Germany's second-largest health insurer, providing a significant representation of the national populace.

The study was approved by the ethics committee of the Jena University Hospital and the data protection officer of the German federal state of Thuringia (the German Clinical Trials Register DRKS00024588). Since claims data were anonymous, the need for informed consent was waived. The description of the study follows the Reporting of studies Conducted using Observational Routinely-collected Data (RECORD) guidelines.[2]

Using the Operationen- und Prozedurenschlüssel (OPS) classification (OPS codes 5-530 or 5-531), which is the German modification of the international classification of procedures in medicine, we identified cases who underwent hernia surgery in 2018. Since CPIP was not coded in the ICD-10, surrogate diagnoses like R10-4 (pelvic and perineum pain; pain with localization in other parts of the lower abdomen; other and unspecified abdominal pain) or M79.25 (Neuralgia and neuritis, unspecified: Pelvic region/thigh) were used. Sociodemographic and health care parameters, such as pain medication, outpatient and multimodal pain therapy, psychiatric comorbidities, physical and occupational therapy were obtained one year before and after the index year 2018.

#### *Patient and control cohorts*

The cross-sectional unicenter *ResolvePAIN* study protocol was registered at the German registry for clinical studies (https://www.germanctr.de/) (Registration Number DRKS00016790). Study participants were recruited from the outpatient clinic of the Center for Interdisciplinary Pain Medicine or approached during a follow-up study at the Dept of Surgery.[3] Patients were $\geq$18 y, of both sexes and gave written informed consent. Ethical approval was obtained from the responsible ethics committees at the study centers. Pain of other origin at the side of investigation, current or past malignancies, surgeries that took place within the past 4 weeks, autoimmune, neurological, and major psychiatric diseases as well as diabetes were exclusion criteria. Seventeen patients were included, 11 males and 6 females (**Table 1**).

The control group of the *ResolvePAIN* study was recruited using newspaper ads and comprised 141 cases based on a sample size calculation using variability of pressure pain thresholds based on age and sex.[4] In addition to the criteria mentioned above, subjects with any type of previous inguinal surgery were excluded from the control group. The controls were subdived by three age groups (**Supplementary Table 1**).

#### *Clinical assessment and patient reported outcomes*

The patient's history included age, education, employment status, medical and drug history as well as family history for chronic pain, psychiatric and neurological diseases. Last menstrual period or if applicable menopausal state were assessed in females. Basic clinical parameters were measured for each participant (weight, height, body mass index). All study participants underwent a clinical neurological examination and quantitative sensory testing.

In addition, patients filled in standardized patient reported outcomes[5]: The neuropathic pain inventory (NPSI), German version, (range 0-100) describes the expression of different neuropathic pain characteristics. It was designed to evaluate the different symptoms of neuropathic pain, to differentiate between subtypes of neuropathic pain and to possibly verify whether symptoms respond differentially to various pharmacological agents or other therapeutic interventions. The Graded Chronic Pain Scale, German version, (GCPS) classifies into four chronic pain grades (Grade I – IV) based on pain intensity items and disability items. Depression was measured by the Beck Depression Inventory 2, German version, (BDI-II, range 0-63). BDI-II results between 0 and 13 are considered minimal depression, 14-19 mild depression, 20-28 moderate depression and 29-63 severe depression. The State-Trait Anxiety Inventory, German Version, (trait anxiety subscale STAI-T, range 20-80) was filled in to record anxiety symptoms. For the STAI-T, a value </= 39 was defined as normal.

#### *Quantitative sensory testing (QST)*

All controls and CPIP patients underwent QST following the standardized DFNS (German Research Network on Neuropathic Pain) protocol.[6] All measurements were performed by a formally trained investigator. The affected inguinal side was defined as the test and the contralateral side as control side. Measurements were conducted halfway between spina iliaca anterior superior and ramus superior of the pubic bone. In control subjects , the thenar region of the hand was assessed first before moving on to the groin.

QST consisted of 11 single tests resulting in 13 parameters, subsequently applied to the selected area beginning with the control side. It includes determination of temperature (cold detection threshold, CDT; warm detection threshold, WDT, thermal sensory limen, TSL) and thermal pain (pain cold pain threshold, CPT; heat pain threshold, HPT) thresholds. Paradoxical heat sensations (i.e., cold stimulation is perceived as heat pain, PHS) were counted if present. Next, mechanical detection threshold (MDT), mechanical pain threshold (MPT), mechanical pain sensitivity (MPS), dynamic mechanical allodynia (DMA), the wind-up ratio for painful stimuli (WUR), the vibration detection threshold (VDT) and the pressure pain threshold (PPT) were determined.

Data from controls were first used to generate normal values grouped by age and sex. Values from CPIP patients were then analyzed after z-transformation. To allow for log-transformation the constant 0.1 was added to the mean value which prevented measurements with a mean of zero from being excluded. After log-transformation the following equation was used to obtain z-values:

$$z = \frac{(value\ of\ the\ subject) - (mean\ of\ healthy\ control\ group)}{SD\ of\ healthy\ control\ group}$$

According to Magerl et al., raw data were used for z-score computation in CPT, WDT and VDT, while other parameters were used in logarithmic form. Z-Scores were calculated with each age- and gender-based control group being used to determine mean and standard deviation needed.[4, 7]

Controls were subdivided into groups according to gender (female, male) and age (18-39 y, 40-59 y, 60-80 y). QST measurements of control subjects' thenar regions were referenced to data previously published by the DFNS (**Supplementary Figure 1**).

#### *Standardized skin biopsies and immunohistochemistry*

Skin biopsies were aseptically obtained using a 5 mm punch after local application of mepivacaine. Due to the small size of the punch, sutures were not needed. The biopsies were performed at the same location as the QST measurements. Bilateral biopsies were obtained from patients and unilateral ones from controls. Each biopsy was divided into two pieces: one part was used for immunohistochemistry, one shock frozen for RNA analysis.

Skin biopsy specimens were fixed in 4% paraformaldehyde for 30 min, then subsequently washed in PBS in three 10 minute-intervals before overnight storage in 10% sucrose until freezing and embedding in OCT. [7, 8] Samples were cut into 50 µm thickness and mounted on microscope slides. The slides were dried at room temperature (RT) for 30 min, outlined with a hydrophobic barrier using Dako Pen, and blocked with 10% BSA for 30 min at RT in a humid chamber. A combination of the primary antibody against PGP9.5 (Zytomed systems, 516-3344, Berlin, Germany, 1:200) overnight at 4°C in a humid chamber and the secondary antibody Cy3 (Jackson ImmunoResearch, West Grove, PA, USA, 1:50) 2 hours at RT before mounting in Vectashield containing DAPI and final storage at 4°C. Nerve fibers were counted according to guideline of the European Federation of Neurological Societies.[8]

#### *Building a prediction model for CPIP*

A predictive model was created to anticipate the presence of CPIP using objective measures. The model was applied to a sample of 17 CPIP patient and 17 age- and sex-matched control groups. For this model, several lab values, QST results, STAI-T, BDI, age and gender were included as predictor variables. Missing values were replaced with the mean for each group and variable, separately. The importance of the selected variables was determined using Random Forests (RF) followed by variable selection using the python package 'scikit-learn'.[9] RF is a machine learning algorithm used for classification and regression, and is widely applied in medical fields.[9] Aside from its ability to provide accurate predictions, RF also incorporates measures for estimating variable importance. One of these measures is the mean decrease in impurity (MDI), which measures variable importance, where higher MDI values are associated with higher importance.[10] Using MDI, we were able to determine the most important five variables to be included in the model. These were Apo A1, HDL, STAI-T, BDNF and CCL2. Prior to entering the model, we found that HDL had high multicoliniarity (VIF >5) because of its correlation with Apo A1, and thus it was removed. The rest of the variables had no multicoliniarity (VIF <2.5). Using the chosen variables as predictors, a binary logistic regression model was built with diagnosis (CPIP vs Control) as the outcome variable, using the python package 'statsmodels'.[11]

### Supplementary tables

| Controls | | | |
|---|---|---|---|
| **Characteristics** | **All (n = 141)** | **Females (n = 68)** | **Males (n = 73)** |
| Age groups | | | |
| 18-39 y (n) | 42 | 21 | 21 |
| 40-59 y (n) | 52 | 23 | 29 |
| 60-80 y (n) | 47 | 24 | 23 |
| BMI [kg/m$^2$] median (range) | 24.1 (19.1-37.4) | 23.7 (19.7-34.3) | 24.3 (20.2-37.4) |
| BDI-II [0-63] median (range) | 2.0 (0 – 24) | 2.0 (0 – 24) | 2.5 (0 – 19) |
| STAI-T [20-80] median (range) | 31.0 (20-54) | 31.0 (20-54) | 32.0 (20-53) |

**Supplementary Table 1: Clinical characteristics of the healthy control group** comparing males and females. BMI, body mass index; BDI-II, Beck Depression Index-II; STAI-T, State-Trait Anxiety Inventory.

| QST Parameter | ρ | p-value |
|---|---|---|
| **CDT** | 0.241 | 0.004 |
| **WDT** | 0.219 | 0.010 |
| **TSL** | 0.259 | 0.002 |
| **CPT** | -0.044 | 0.607 |
| **HPT** | 0.034 | 0.696 |
| **MDT** | 0.198 | 0.020 |
| **MPT** | 0.193 | 0.023 |
| **MPS** | -0.160 | 0.060 |
| **WUR** | 0.45 | 0.609 |
| **VDT** | -0.182 | 0.032 |
| **PPT** | 0.162 | 0.058 |

**Supplementary Table 2: Spearman's correlation between BMI and QST parameters.** BMI, body mass index; QST, quantitative sensory testing; CDT, cold detection threshold; WDT, warm detection threshold; TSL, thermal sensory limen; HPT, heat pain threshold; CPT, cold pain threshold; MDT, mechanical detection threshold; MPS, mechanical pain sensitivity; MPT, mechanical pain threshold; WUR, wind-up ratio; VDT, vibration detection threshold; PPT, pain pressure threshold.

| CPIP patients | | |
|---|---|---|
| **Characteristics** | **Pain persistence (n = 7)** | **Novel pain (n = 7)** |
| Age at inclusion [years] median (range) | 42 (33 - 67) | 48 (25 - 73) |
| Age at operation [years] median (range) | 40 (30 - 62) | 50 (23 - 70) |

| | | |
|---|---|---|
| Years since surgery median (range) | 3 (1 - 5) | 2 (1 - 6) |
| BMI [kg/m$^2$] median (range) | 24 (21 - 28) | 23 (18 - 30) |
| **Type of surgery (number)** | | |
| Open/TEP/TAP/not specified | 4/2/1/0 | 2/2/1/2 |
| **Pain & mental health assessment** | | |
| Mean pain [0-10] median (range) | 4 (4 - 5) | 3 (2 - 5) |
| Min. pain [0-10] median (range) | 2 (1 - 6) | 2 (0 - 5) |
| Max. pain [0-10] median (range) | 8 (4 - 10) | 6 (3 - 10) |
| NPSI [0-100] median (range) | 20 (2 - 32) | 14 (0 - 61) |
| **GCPS [I-IV]** (n) | | |
| **I/II/III/IV** | 3/4/0/0 | 4/1/0/0 |
| BDI-II [0-63] median (range) | 8 (0 - 18) | 2 (0 - 19) |
| BDI-II patients over cut-off mild depression (n) | 3 | 2 |
| STAI-T [20-80] median (range) | 36 (20 - 48) | 30 (26 - 39) |
| STAI-T patients over cut-off for anxiety (n) | 2 | 0 |
| **Treatment (n-number)** | | |
| Non-Opioids/Opioids/Antineuropathics | 4/1/2 | 3/1/0 |
| Local treatment | 1 | 0 |
| Surgical neurolysis | 0 | 0 |
| Physiotherapy | 2 | 3 |

**Supplementary Table 3: Clinical characteristics of CPIP patients comparing with pain persistence (pain pre surgery) and new pain (no pain before surgery).** BMI, body mass index; TEP, total extraperitoneal patch plasty; TAPP, transabdominal preperitoneal patch plasty; NPSI, Neuropathic Pain Symptom Inventory; GCPS, Graded Chronic Pain Scale; BDI-II, Beck's Depression Index-II; STAI-T, State-Trait Anxiety Inventory-Trait. In case numbers did not add up to 14, items were not available for all patients.
